# Metagenomics - AI powered prediction of Inflammatory Bowel’s Disease and Probiotic Recommendation

**DOI:** 10.64898/2026.02.12.26345333

**Authors:** Sastha KumarN, Mathew Thomas, Sai Janakiram Chinnakanu, M Nidheesh, Sabarinath Subramaniam

## Abstract

**Background and Objective:** The dysbiosis of human gut microbiome has been increasingly seen to have a relation in the development of autoimmune diseases, with specific microbial signatures having causative association with specific conditions. Inflammatory bowel disease (IBD) is one such autoimmune ailment. This paper proposes a predictive tool that can identify the IBD status of an individual based on the composition of the gut microbiome using machine learning and AI agents driven techniques. The technology can strengthen the suspicion of a potential IBD diagnosis a patient may have based on their gut microbiome profile.

**Methods:** The tool processes patient gut metagenome using integrated Kneaddata and MetaPhlAn to generate taxonomic profiles. These are fed into an XGBoost classifier to predict IBD or healthy status. Dysbiotic taxa are identified via Z-score and fold change. CrewAI delivers personalized probiotic recommendations based on diagnosis and dysbiosis.

**Results:** The tuned XGBoost model achieved 86.6% accuracy. On validation using single ulcerative colitis sample, the tool correctly predicted IBD status but misclassified it as Crohn’s disease(possibly due to overlapping microbial signatures), identifying Faecalibacterium and Flavonifractor as dysbiotic taxa.The probiotic recommended was Faecalibacterium prausnitzii, backed with reasoning basedon scientific literature.

**Conclusions:** Despite limited validation sample size, the high accuracy , correct IBD detection,dysbiosis analysis and elaborate probiotic recommendation suggest promising potential; further validation needed

## 1. Introduction

The human body consists of a vast community of trillions of microbes, collectively known as the human microbiome, with the gastrointestinal tract, particularly the colon, being the most densely populated habitat collectively called the human gut microbiome. This intricate community, composed of over 1000 bacterial species, maintains a crucial symbiotic relationship with the host, influencing vital processes such as digestion, vitamin production, and immune system modulation [1].While the taxonomic profiles of individual microbiomes vary significantly due to factors like geography, diet, and host genetics, their underlying functional potential via the metabolic pathways shows remarkable consistency across populations. However, any imbalance in the composition and function of this microbiome, which is consistent, is referred to as dysbiosis. Dysbiosis is strongly linked to the development and progression of chronic conditions, including metabolic, autoimmune, and neurological diseases. Currently, it is increasingly recognized as a key environmental factor that can shift the immune response towards pro-inflammatory pathways, thereby instigating autoimmune processes[2].

IBDs, encompassing Crohn’s disease (CD) and Ulcerative Colitis (UC), are characterized by chronic gastrointestinal inflammation and exhibit altered gut microbiome compositions, including reduced bacterial diversity, a decrease in beneficial bacteria (like *Faecalibacterium prausnitzii*) , [3]and increase in potentially harmful or opportunistic taxa (like Proteobacteria, *Escherichia-Shigella*, and *Enterococcus*)[4].

The mechanisms by which gut dysbiosis contributes to autoimmune diseases include the loss of intestinal barrier integrity, which is popularly referred to as “Leaky Gut,” and has been observed in various autoimmune diseases. Tight junctions, which include proteins like occludin, are crucial for maintaining the barrier function. Disruption of the barrier allows microbial products and bacteria to leak into the host tissues and bloodstream, potentially causing the immune system to respond [5,6]. This allows microbial products to trigger systemic inflammation and molecular mimicry, where microbial antigens structurally similar to some host self-antigens result in the onset of an immune reaction against the self-antigens. Also, the modulation of the immune system by microbial metabolites like short-chain fatty acids (SCFAs) can lead to an increase in systemic inflammation[7].

According to current research, diagnostic delays are frequent; on average, they occur between 2 months and 8 years after the onset of symptoms [8].The diagnosis delay for UC is usually 4 months, while for CD, it is roughly 9 months. Children and mild or vague presentation cases are more likely to have delays [9].Traditional IBD diagnostic methods face challenges due to symptom variability and invasiveness, which can hinder an accurate and timely diagnosis. The validity and reproducibility of evaluations are affected by the significant variability between observers that hinders endoscopic scoring systems as a whole, such as the Mayo score, which is utilized for assessing the progression of the disease[10].

In response, machine learning (ML), combined with high-throughput sequencing technologies like shotgun metagenomics, has emerged as a powerful computational approach for analyzing complex microbiome data, enabling the accurate classification of IBD patients and the identification of microbial biomarkers for disease prediction [11].

Improvements in metagenomics, metatranscriptomics, and metabolomics have become essential in deciphering the intricate interactions between the gut microbiome and IBD. Integration of metagenomics data with host genomic data and clinical metadata is crucial to deciphering the intricate host-microbe interactions underlying IBD and designing microbiome-based diagnostics, prognostics, and therapeutic interventions.

Probiotics are live microorganisms that, when administered adequately, confer health benefits through various mechanisms, including direct antimicrobial activity, strengthening the intestinal barrier by enhancing tight junctions, and modulating the host immune system to reduce inflammation[12,13]. While evidence for their effectiveness in Crohn’s disease is often limited, substantial evidence supports use of specific probiotic strains, such as *Escherichia coli Nissle 1917* and the multi-strain preparation VSL#3, for inducing and maintaining remission in patients with mild to moderate Ulcerative Colitis [14]. While generally considered safe one must use only specific probiotic strains which are documented to have beneficial effects on mild or moderate inflammatory diseases, as wrong recommendations could result in side effects in immunocompromised patients. Crucially, the efficacy of probiotics is strain-specific and dose-dependent, hence the variability in research findings [15].This highlights the need for personalized insights on what probiotic strains could be useful for a particular individual [16].This paper presents a Metagenomic-ML-based tool that predicts IBD via gut microbiome profile and uses CrewAI to deliver personalized probiotics recommendation.

## 2. Methodology

### 2.1 Study Design

The tool aims to fulfill the following purpose: predicting user’s disease state and, if diseased, recommending suitable probiotic. The tool is structured into four distinct parts: generating a user’s taxonomic profile, predicting the disease using a machine learning model that is trained on the taxonomic profiles of healthy individuals and individuals with IBD, identifying the dysbiotic taxa, and thereby providing personalized probiotic recommendations using CrewAI, if a disease state exists.

### 2.2 Data Collection

#### 2.2.1 Dataset for IBD prediction model

The data utilized for this project were derived from shotgun metagenomic sequencing, which provides detailed taxonomic profiles of microbial communities [17].Two primary sources were integrated: Dataset 1 was retrieved from the Inflammatory Bowel Disease Multi’omics Database (<u>Dataset 1</u>), part of the Integrative Human Microbiome Project (HMP2), contributing 1638 samples with taxonomic classification up to the strain level. Dataset 2 originated from the results of the paper “Gut microbiome structure and metabolic activity in inflammatory bowel disease” [18], comprising 220 stool samples from two cohorts (155 from Massachusetts, 65 from the Netherlands). These samples were from individuals with Crohn’s disease (CD), Ulcerative colitis (UC), and non-IBD controls, with taxonomic classification up to the species level (<u>Dataset 2</u>). Both the dataset included metadata features Age, Immunosuppressant usage, Antibiotic usage, and Diagnosis.

#### 2.2.2 Dataset for metagenomic tools

Tools like Knead Data and MetaPhlAn2 were used to process the users’ metagenome, with KneadData using the GRCh38 human genome to remove host DNA and MetaPhlAn using the mpa_v31_CHOCOPhlAn_201901 database for taxonomic profiling based on over 17,000 microbial genomes. Both these tools were incorporated into our pipeline.

### 2.3 Data Preprocessing

A significant challenge in integrating these datasets was their inconsistency, specifically differing levels of taxonomic classification. To address this, a preprocessing approach was employed, involving two key steps: First, collapsing the organisms to a higher taxonomic level, specifically the genus level, by summing the relative abundances of species or strains within the same genus . This increased feature commonality between datasets. Second, the datasets were merged, ensuring that all features (organisms and metadata) from both datasets were retained, with uncommon taxa assigned a relative abundance of zero to maintain sum totals. The resulting merged dataset contained 1858 samples and a taxonomic profile of 195 genera, representing 485 healthy, 838 CD, and 535 UC samples. Other preprocessing steps included relative abundance normalization (renormalizing genus-level abundances to sum to 1), metadata encoding (e.g., Diagnosis: Healthy=0, CD=1, UC=2; Immunosuppressant/Antibiotic Usage: Yes=1, No=0), and min-max normalization of Age to a range between 0 and 1.

### 2.4 Tool Development and Evaluation

#### 2.4.1 User Taxonomic Profile Generation

The first part of the methodology focuses on User Taxonomic Profile Generation. This begins with processing raw paired-end FASTQ files from the user’s gut metagenome sample. Kneaddata (v0.12.0) is employed to meticulously remove host-contaminant DNA, specifically human DNA, and low-quality reads . This step involves several integrated tools: Bowtie2 (v2.4.1) is used for alignment against the GRCh38 human genome (a pre-indexed database), and Trimmomatic (v0.39) handles adapter trimming and quality filtering [19].Following host DNA removal, these processed paired-end reads are fed into MetaPhlAn3 (v3.0.14) to generate the crucial taxonomic profile. MetaPhlAn3 utilizes the mpa_v31_CHOCOPhlAn_201901 database for its profiling, specifically designed to output genus-level abundances by using the --tax_lev g flag [20]. The relative abundances of these genera are then normalized to sum to 1.0 per sample. Unclassified genera are removed, and the profile undergoes feature alignment for consistency with training dataset, making it a valid input for prediction [21].

#### 2.4.2 Disease Prediction Using XGBoost

The second core component is Disease Prediction, which leverages an XGBoost machine learning model. XGBoost was strategically chosen due to its robust capabilities in handling high-dimensional data typical of gut microbiome profiles (such as 195 genera and 4 clinical metadata features) [22]. Its effectiveness stems from its ability to model the complex and nonlinear patterns and interactions among genera that determine an individual’s IBD status [23].

The model’s hyperparameters are carefully tuned to ensure optimal performance. To find the best combination of hyperparameters, RandomSearchCV is employed. This method randomly samples 20 combinations from the defined parameter space, evaluating each using 5-fold cross-validation with accuracy as the scoring metric. This approach efficiently identifies optimal settings, striking a balance between model complexity and generalization capabilities, thereby enhancing prediction accuracy [24].

The XGBoost estimator is configured to use multi-class logarithmic loss to rank feature importance. Recursive feature elimination is used to iteratively remove the lowest-ranked features until a predefined number of 180 genera are selected. This feature selection is performed exclusively on the training dataset, which is an 80/20 split of the main dataset, to prevent information leakage into the test set. The selected genera are then consistently applied to the test dataset. The XGBoost model itself is optimized for multi-class logarithmic loss, which is ideal for the CD/UC/Healthy classification task.

#### 2.4.3 Dysbiotic Taxa Prediction

The third critical component is the dysbiotic taxa identification.This tool identifies the top 2 dysbiotic taxa that are most altered compared to a healthy gut microbiome and are likely contributing to the inflammatory environment [25]. To achieve this, a healthy cohort baseline dataset is established by filtering all healthy samples from the same dataset used to train the XGBoost model and then taking the average of the relative abundances of each genus present in each healthy sample, thereby creating a baseline healthy taxonomic profile. This baseline is then used for comparison against the patient’s taxonomic profile.

Two statistical techniques are used for Dysbiosis Quantification: Z-score and Fold Change Z-score measures how many standard deviations away a patient’s genus abundance is from the mean abundance of that genus in the healthy baseline.The formulae is shown in equation (1) in Figure 1 where X is the observed abundance, µ is the mean healthy abundance, and σ is the standard deviation of healthy abundances. Genera with a Z-score greater than 1.5 are considered to have a significant deviation and are ranked in descending order.

Fold Change measures the relative difference between the patient’s genus abundance and the mean healthy abundance .The formula is shown in equation (2) in Figure 1 where X is the patient’s abundance, µ is the mean healthy abundance, and a small pseudocount (1e-6) is added to avoid division by zero.Genera with a fold change > 1.5 (enriched) or < 0.67 (depleted) are considered dysbiotic and are also ranked.The top two dysbiotic taxa are selected primarily based on their Z-score ranking. Fold change serves as a secondary ranking criterion, acting as a tiebreaker when Z-scores are similar. Taxa with zero abundances in both healthy samples and the patient’s profile are disregarded.

#### 2.4.4 Probiotic Recommendation

The Probiotic Recommendation System utilizes a CrewAI setup [26], which is a multi-AI agent framework designed to automate personalized and precise probiotic recommendations

This setup features two main agents:

The Microbiome Research Scientist agent conducts in-depth literature reviews, leveraging current scientific articles and clinical guidelines related to probiotics based on the provided disease condition and dysbiotic taxa.

The Clinical Probiotic Specialist agent analyzes the findings from the research scientist agent and recommends the most biologically appropriate and clinically applicable probiotic for the patient, providing an explanation of its mechanism of action, particularly on the identified dysbiotic taxa

The interaction between these agents is coordinated by a Crew object.The system requires three specific inputs from the user: the Disease name (e.g., Crohn’s Disease), Dysbiotic Taxa 1 genus (e.g., Klebsiella), and Dysbiotic Taxa 2 genus (e.g., Salmonella). Both agents are equipped with TavilySearchTool, an internet-based API integrated into the Crew pipeline, enabling them to access recent peer-reviewed literature, probiotic case studies, and clinical guidelines. This internet access is crucial for ensuring the recommendations are precise, accurate, and clinically relevant based on the most current research. The agents’ reasoning and recommendation generation are powered by Gemma 3 [27], a large language model that is installed locally. The prompts for these agents are meticulously engineered, defining their specific Role and Goal, providing a Backstory to contextualize their domain, and offering Task Instructions with a clear schema for their expected output. This structured approach ensures the generated recommendations are clear, consistent, and easily interpretable.

### 2.5 Tool Interface integration

We use the Streamlit interface to integrate the two components, the dysbiotic taxa prediction pipeline and the probiotic recommendation pipeline, to form a user-friendly and intuitive tool that users can use to enter their gut metagenome and receive the dysbiosis analysis and the probiotic recommendation.

## 3. Results

### 3.1 XGBoost Model Results

#### 3.1.1 RandomSearchCV results

RandomSearchCV was used to tune the hyperparameters of the model. The best hyperparameters obtained for the model include a subsample of 0.8, meaning 80% of the training data is used per tree, and a reg_lambda of 1, which applies moderate regularization to balance bias and variance. The model uses 200 trees (n_estimators) to capture complex patterns, with a max_depth of 10 allowing deep trees, though regularization and subsampling mitigate overfitting risks. A learning rate of 0.2 ensures significant contributions from each tree, supported by regularization to prevent overfitting. A gamma of 0.1 ensures splits only occur with a minimum loss reduction, avoiding unnecessary complexity, and a colsample_bytree of 1 indicates all features are used, suitable when all are relevant for prediction.

#### 3.1.2 XGBoost Model Results

The XGBoost model got an overall Accuracy of 86.56%, showing strong performance in classifying samples as CD, UC, or Healthy, as shown in Figure 2. The model excelled in identifying UC, with a Precision of 0.9082, a Recall of 0.8990, and F1 Score of 0.9036, suggesting high reliability for UC diagnosis.

For CD, the model showed a high recall of 93.98 %, ensuring most CD cases were correctly identified, though its precision of 84.32% indicates some false positives, as the F1 Score is 88.89 %. However, performance on the Healthy class was weaker, with a Recall of 71.96 % and an F1 Score of 078.57 %.The confusion matrix shown in Figure 3 shows strong UC performance, mostly satisfactory CD (minor misclassifications), but notable Healthy to CD misclassification, yielding Recall: 71.96%, F1: 78.57%, and Precision: 86.52% for Healthy.

Given in Figure 4, we can see the top 10 features contributing the most to the prediction. The immunosuppressant metadata feature contributes the most to prediction. Inflammatory diseases, like IBD, are autoimmune in nature . Immunosuppressants reduce the strength of the immune system, thereby reducing autoimmune symptoms as well. Patient response to immunosuppressants is also used by physicians as a way to diagnose IBD [28]. This features overcontribution can explain the misclassification of “Healthy” groups (Non-IBD) as CD.

### 3.2 Validation

For validation, the sample CSM9X233 (human gut metagenome) from the Inflammatory Bowel Disease Multi-omics Database (IBDMDB), part of the longitudinal multi-omics study “Human microbiome in Inflammatory Bowel Disease” was selected . The sample was sequenced using the Illumina HiSeq 2000 platform with whole-genome shotgun metagenomics (paired-end), yielding run SRR5950521 (∼9.65 million reads, 626.7 MB compressed file size). The data were submitted by Harvard T.H. Chan School of Public Health and made publicly available on 2017-08-22.

The sample is taken from a Crohn’s disease(CD) patient aged 45 years old with antibiotic and immunosuppressant usage.The fastq files (forward and reverse reads)are uploaded to the tool.The metagenomic processing gives us a taxonomic profile of the patient. This prepared profile is then used for XGBoost prediction.

#### 3.2.1 XGBoost model Output

We feed the input to the XGBoost model. The user is instructed to enter the metadata as we mentioned before. We enter the following details for metadata.The model prediction shown in Figure 5 shows the diagnosis as Crohn’s Disease , which is a misdiagnosis of the single Ulcerative Colitis patient sample.Since the validation sample size is very small the tool still shows potential as seen in performance metrics ( Accuracy: 86.56%); further validation is required.

#### 3.2.2 Dysbiosis analysis Output

Using Z score and fold change, the top 2 dysbiotic taxa or genera are found to be Faecalibacterium and Flavonifractor [29].Table 2 shows that Faecalibacterium and Flavonifractor pass the criteria and are the top two in rank for being dysbiotic, considering both z-score and fold change.

**Table 1:** Best hyperparameter obtained through RandomizedSearchCV.

| Hyperparameter | Value |
| --- | --- |
| subsample | 0.8 |
| reg_lambda | 1 |
| n_estimators | 200 |
| max_depth | 10 |
| learning_rate | 0.2 |
| gamma | 0.1 |
| colsample_bytree | 1.0 |

**Table 2:** The table shows the Z- and fold change of the top 2 dysbiotic taxa. Faecalibacterium and Flavonifractor are two taxa or genera that cause.

| Genus | Z-score | Fold Change | Dysbiotic | Z-score Rank | Fold Change Rank |
| --- | --- | --- | --- | --- | --- |
| Faecalibacterium | 3.107 | 4.05 | True | 1 | 4 |
| Flavonifractor | 1.63 | 5.13 | True | 2 | 1 |

#### 3.2.3 Crew AI Probiotic Recommendation Output

The dysbiotic taxa and the predicted disease information is given to Crew AI to get the result.The recommendation by Crew AI setup is Faecalibacterium prausnitzii with the mechanistic reasoning on why it is recommended based on its action on the gut as shown Figure 6.This recommendation is for assistive purposes only. To give user an insight into their health, and should not be taken as medical advice.

## 4. Discussion

The methodology and results of this paper provide a robust framework for predicting inflammatory bowel disease (IBD) states and recommending probiotics using a multi-step pipeline that integrates metagenomic data processing, machine learning, dysbiosis analysis, and AI agent-driven recommendations. The pipeline successfully achieves its dual goals of disease prediction and probiotic recommendation, but there are several key aspects to discuss, including the strengths of the approach, limitations, clinical implications, and avenues for future improvement.

The computational pipeline excels in processing shotgun metagenomic data by integrating KneadData(for host DNA removal via GRCh38) and MetaPhlAn (genus-level profiling using mpa_v31_CHOCOPhlAn_201901) with machine learning. It resolves taxonomic inconsistencies across datasets by collapsing to genus level and imputing zero-abundance for missing taxa, yielding a unified training set of 1,858 samples with 195 genera and four metadata features. This ensures robust, consistent input for the XGBoost model.

The XGBoost model, tuned via RandomSearchCV, achieves 86.56% accuracy, with strong performance in UC (Precision: 0.91, Recall: 0.90, F1: 0.90) and CD (Recall: 0.94).It effectively handles high-dimensional taxonomic data (195 genera) and complex interactions, with tuned hyperparameters(max_depth=10,learning_rate=0.2,subsample=0.8,reg_lambda=1,gamma=0.1) balancing complexity and generalization to prevent overfitting.High recall for CD and UC minimizes false negatives, critical for timely clinical intervention

The dysbiosis analysis, using Z-scores and fold changes, identifies the top dysbiotic genera, providing a data-driven basis for probiotic recommendation, although this method is not standard but it is conceptually sound enough. The Crew AI setup, powered by the Gemma 3 LLM and Tavily search, adds a layer of interpretability and clinical relevance by recommending probiotics with explanations of their mechanisms of action. This integration of machine learning with AI-driven recommendation systems is a significant step toward personalized probiotics, as it tailors interventions to the patient’s specific disease state and dysbiotic taxa.

Despite its strengths, the pipeline has notable limitations. The pipeline’s main limitation is the misdiagnosis of single validation Ulcerative Colitis sample as Crohn’s Disease despite predicting the IBD status correctly, this could possibly be due to overlapping of microbial profile in UC and CD.The limited validation sample size suggests further validation is needed. Poor Healthy class performance (Recall: 0.72, F1: 0.79) due to class imbalance (485 Healthy vs. 839 CD, 536 UC). This imbalance likely contributes to the misclassification of Healthy samples as CD., This biases the model toward over-predicting CD/UC to improve overall accuracy, especially influenced by the high-weighted “Immunosuppressants” feature, causing false positives in Healthy patients on such drugs for other inflammatory conditions. Clinically, this risks unnecessary testing, treatment, anxiety, and costs.

The Crew AI setup depends on scientific literature accessed via Tavily. If recent studies or clinical guidelines on probiotics for specific taxa (e.g., Faecalibacterium, Flavonifractor) are limited, the recommendations may lack depth or accuracy. Nevertheless the tool still has potential to improve upon these limitations with more high quality training data and validation samples.

## 5. Conclusion

The pipeline’s ability to predict IBD with high sensitivity (Recall for CD: 0.9398, UC: 0.8990) is promising for clinical applications, as it minimizes the risk of missing diseased patients who need timely intervention. However, the misclassification of Healthy samples as CD or UC (low Recall for Healthy: 0.7196) poses a challenge for clinical adoption, as overdiagnosis could lead to unnecessary procedures. Addressing class imbalance through techniques like SMOTE oversampling or adjusting class weights in XGBoost (e.g., increasing the weight for Healthy samples) could improve performance on the Healthy class, reducing false positives.

The identification of dysbiotic taxa like Faecalibacterium and Flavonifractor aligns with existing literature, as Faecalibacterium (specifically Faecalibacterium prausnitzii) is often depleted in IBD patients and associated with anti-inflammatory properties . The recommendation of Faecalibacterium prausnitzii by Crew AI is plausible, as this probiotic is a key anti-inflammatory commensal bacterium . However, clinical validation through trials is necessary to confirm the efficacy of these recommendations, as the tool’s outputs are assistive and not a substitute for medical advice.

Future improvements could include incorporating species- or strain-level taxonomic profiling to enhance the precision of dysbiosis analysis and probiotic recommendations. Expanding the Healthy baseline dataset to include diverse populations would improve the generalizability of the dysbiosis analysis. Additionally, integrating more metadata features (e.g., dietary habits, disease severity) could improve the XGBoost model’s performance and provide a more holistic view of the patient’s condition. Finally, validating the Crew AI recommendations against clinical outcomes in IBD patients would strengthen the tool’s utility in personalized medicine.

## Data Availability

All data produced in the present study are available upon reasonable request to the authors

## Acknowledgements

This work was carried out at GenAI Research Labs, an initiative of Sivasakthi Science Foundation. The authors gratefully acknowledge the guidance and supervision of Dr. Sabarinath Subramaniam, PhD, whose expertise, insightful feedback, and continuous support were instrumental throughout the project.

## Ethics Statement

This study utilized anonymized, publicly available datasets. No ethical approval was required.

## Declaration of generative AI and AI-assisted technologies in the manuscript preparation process

During the preparation of this work the authors used Notebooklm and Perplexity in order to gain insight from/summarize research papers and finding references respectively. After using this tool/service, the authors reviewed and edited the content as needed and takes full responsibility for the content of the published article.

---

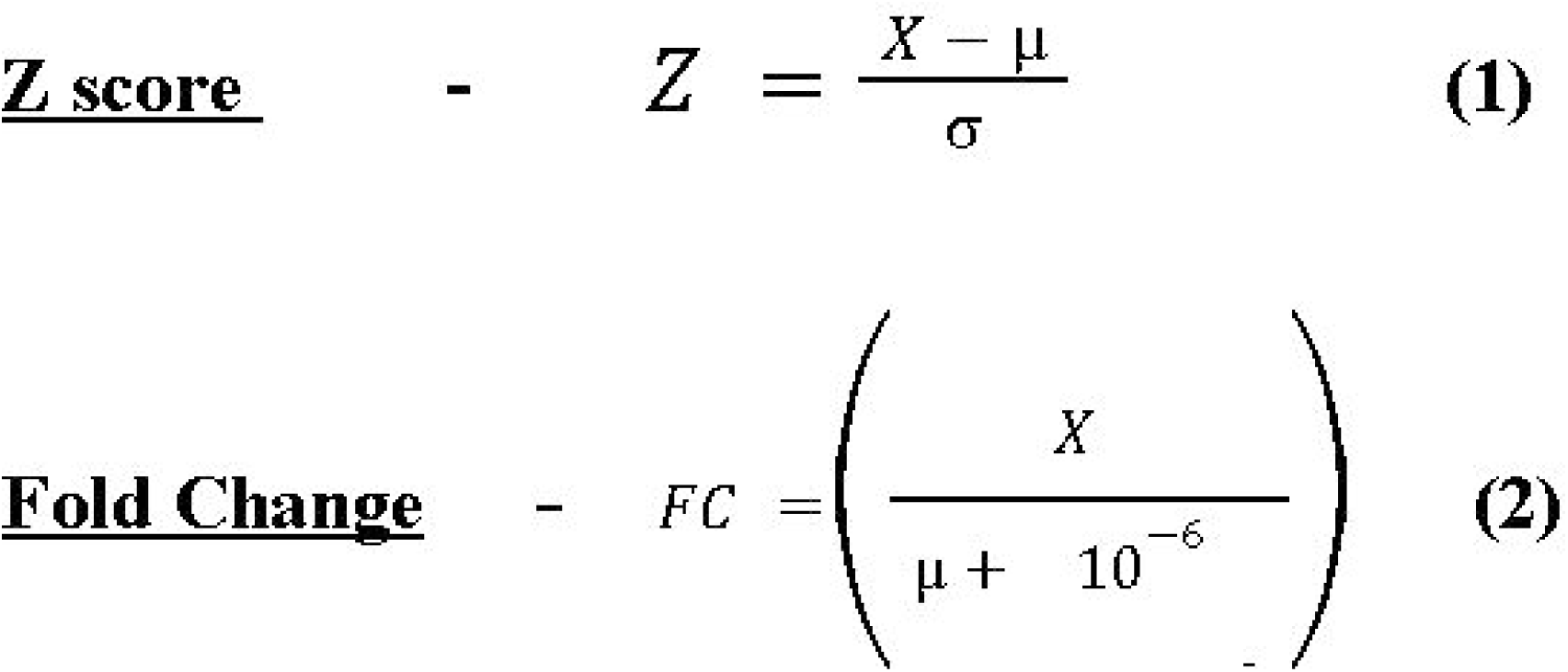

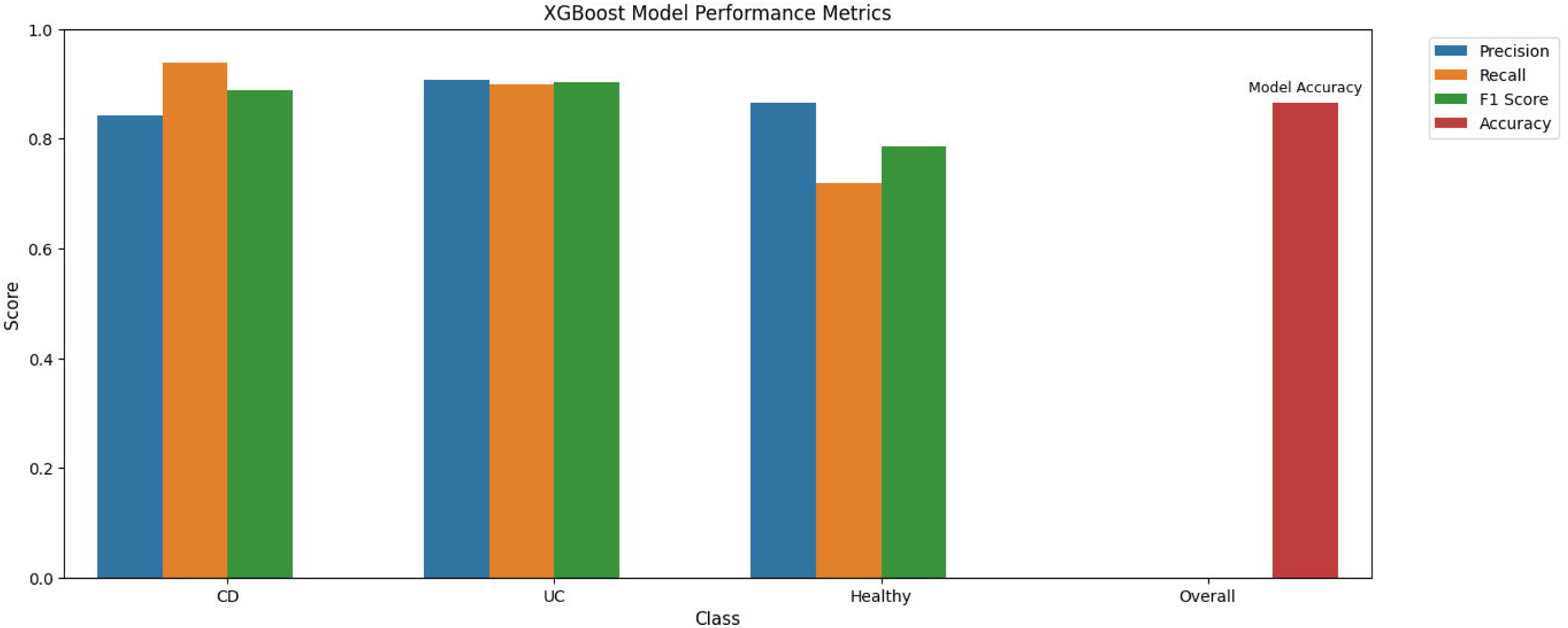

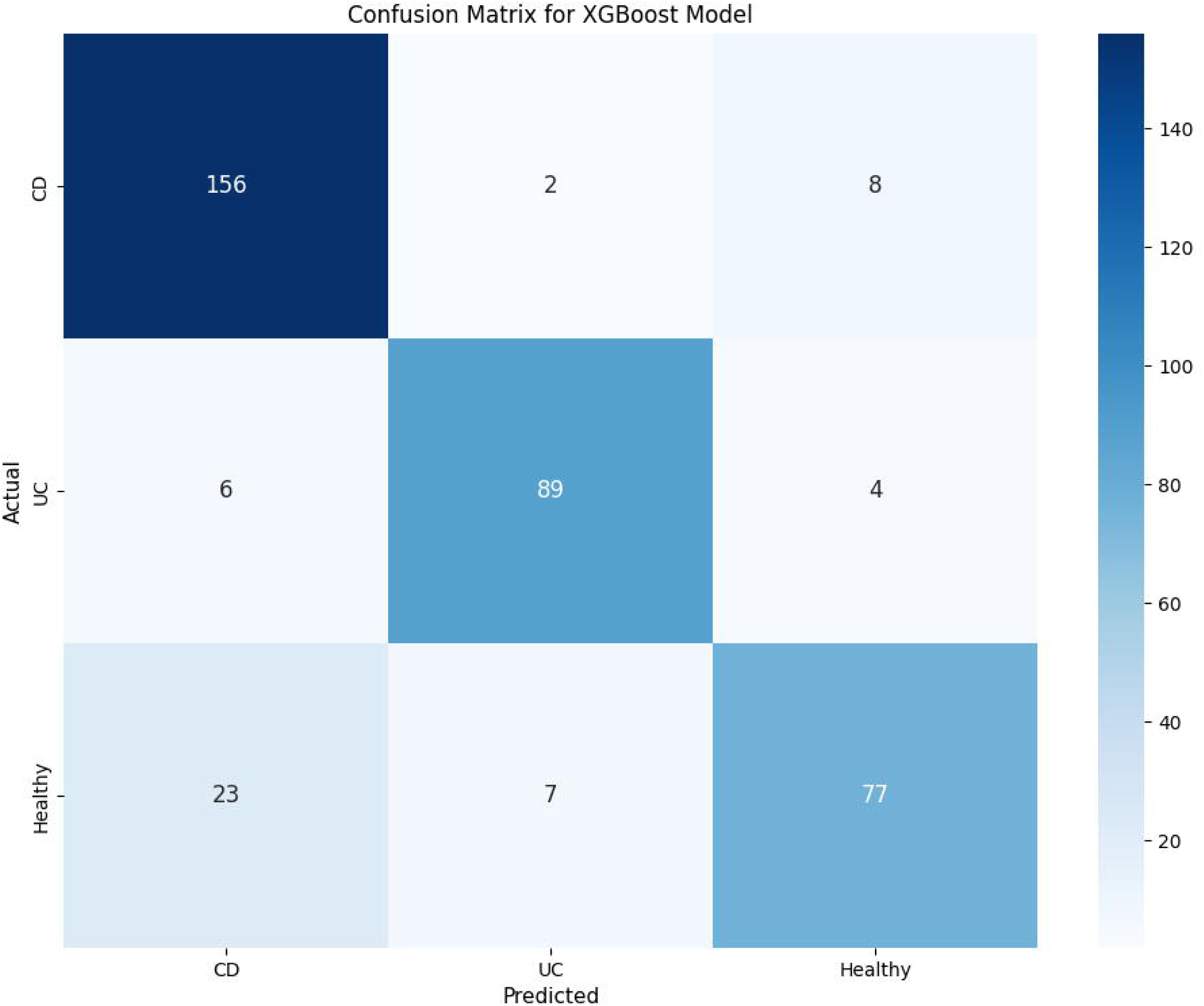

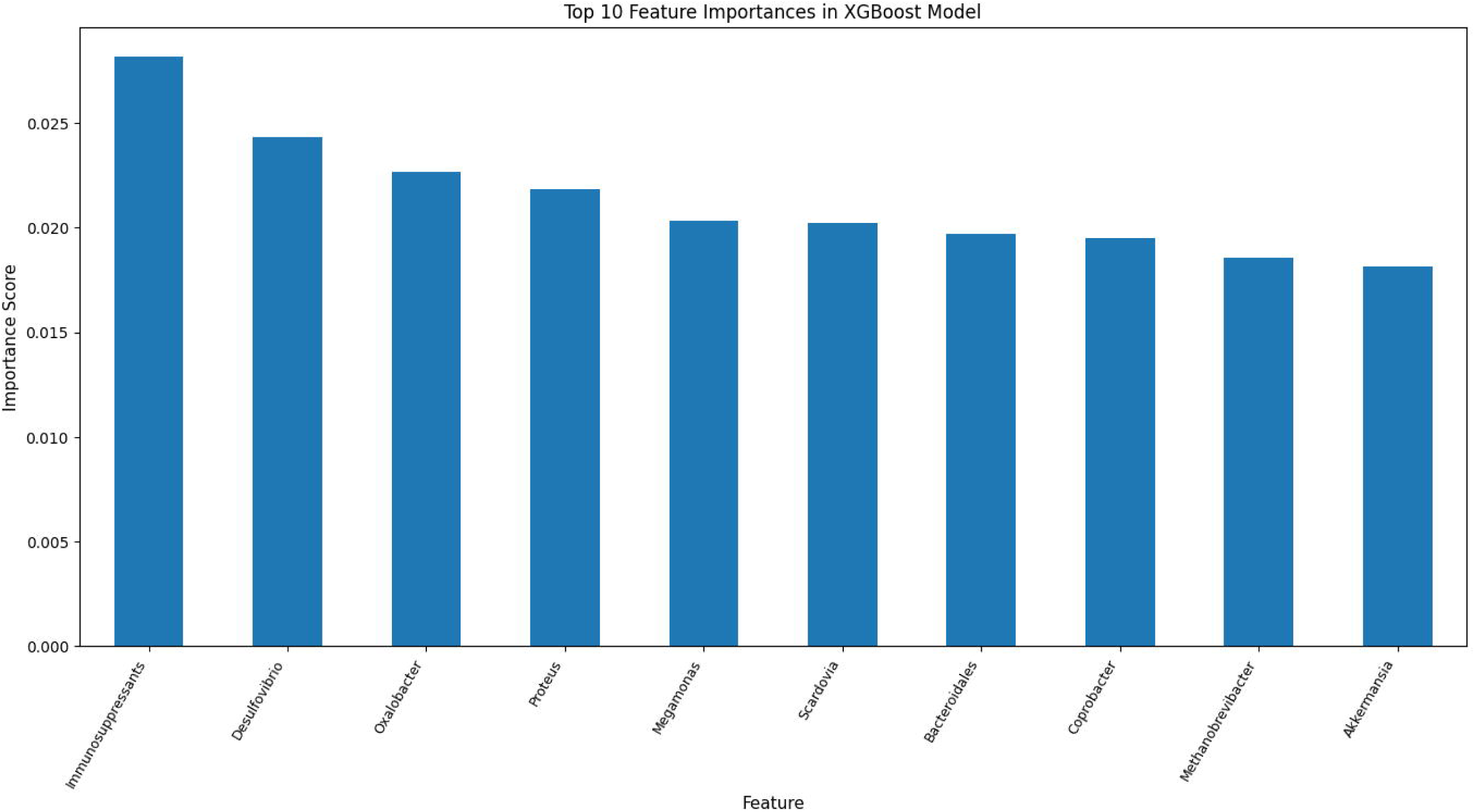

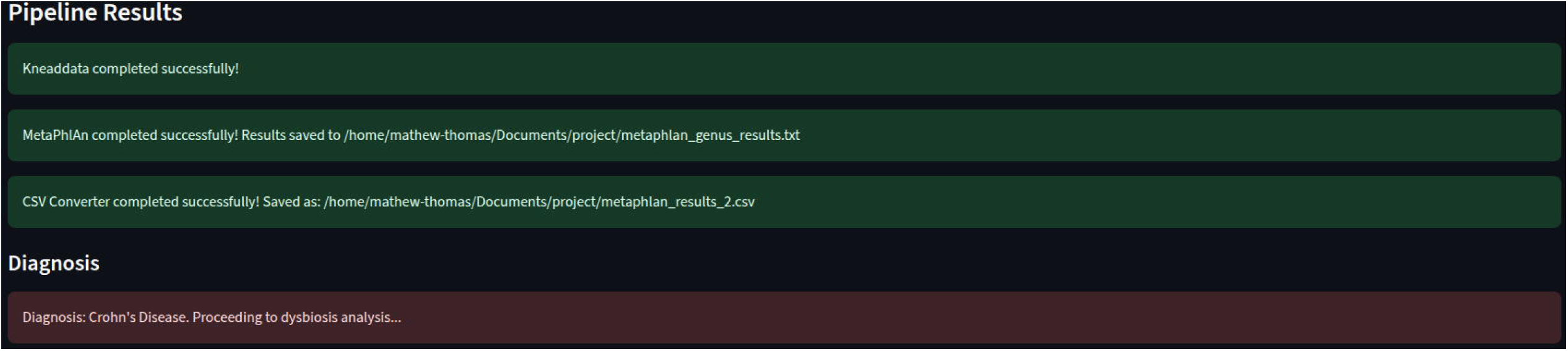

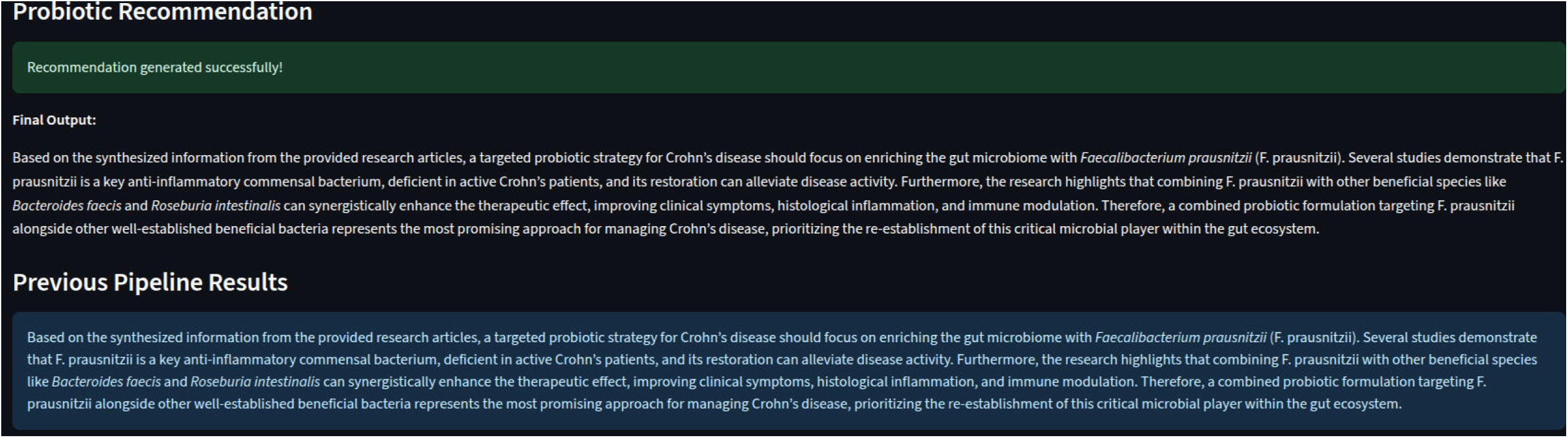

